# DNAJC12 in monoamine metabolism, neurodevelopment and neurodegeneration

**DOI:** 10.1101/2023.06.22.23291747

**Authors:** Isaac Bul Deng, Jordan Follett, Mengfei Bu, Matthew J. Farrer

## Abstract

Recent studies show that mutations in *DNAJC12*, a co-chaperone for monoamine synthesis may cause mild hyperphenylalaninemia with infantile dystonia, young-onset parkinsonism, developmental delay and cognitive deficits. To this end, *DNAJC12* gene has been included in newborn screening, most revealingly in Spain, and those results are a testament to the importance of early diagnosis and treatment in combating human diseases. However, practitioners may be unaware of these advances and it is probable that many patients, especially adults, have yet to receive molecular testing for *DNAJC12*. Therefore, this review summarizes genotype-phenotype relationships and treatment paradigms for patients with *DNAJC12* mutations. It provides an overview of the structure of DNAJC12 protein, known mutations, domains and binding partners, and elaborates on its role in monoamine synthesis, disease etiology and pathogenesis.

## Introduction

Molecular chaperones are molecularly and functionally diverse families of proteins essential for protein folding, trafficking and degradation.^1,2^ Often termed heat shock proteins (HSPs), these include HSP90, HSP70, HSP60, HSP40 and the small HSPs. HSP40 proteins, also known as DNAJ proteins, are further divided into three classes: DNAJAs, DNAJBs and DNAJCs, based on the composition of their domains.^1,2^ DNAJ-containing proteins bind to specific client proteins and HSP-70, synergistically positioning them in proximity to the J-domain to stimulate the ATPase activity of HSP-70, in turn liberating both ADP and folded substrate from this reaction.^2^ HSP-70 protects client proteins’ hydrophobic ends, facilitating folding and preventing protein aggregation.^2,3^ In addition, HSP-70 chaperones target proteins implicated in neurodegeneration including alpha-synuclein and tau for degradation by chaperone-mediated autophagy.^4–6^ Thus, DNAJ-domain containing proteins are necessary to maintain the cellular proteome.

Recent studies show that biallelic recessively-inherited mutations in *DNAJC12*, a member of the DNAJC family of co-chaperones, result in a multitude of neurological symptoms, including developmental delay, intellectual disability, neuropsychiatric disorders, infantile dystonia or young-onset parkinsonism. Mutations in *DNAJC12* also account for a small proportion of hyperphenylalaninemia (HPA) cases that are not attributed to mutations in phenylalanine hydroxylase (PAH) or synthesis of its co-factor tetrahydrobiopterin (BH_4_).^7–9^ The role of DNAJC12 protein is not fully understood, but it appears to function as a co-chaperone for aromatic amino acid hydroxylases (AAAHs) including PAH, tyrosine hydroxylase (TH) and tryptophan hydroxylase 1 and 2 (TPH1 and TPH2), which are essential for the synthesis of dopamine and serotonin. These neurotransmitters are markedly reduced in patients with *DNAJC12* mutations.^7,8^ *DNAJC12* mutations were first discovered in children with HPA, neurodevelopmental delay and dystonia ^7^, and in patients and families with young-onset parkinsonism ^8^, and were quickly appreciated to be important in the differential diagnosis of HPA.^10^ DNAJC12 is the fifth DNAJC family member to be implicated in parkinsonism, with others including DNAJC5 (cysteine string protein-alpha), DNAJC6 (auxillin), DNAJC13 (receptor-mediated endocytosis-8) and DNAJC26 (cyclin G associated kinase (GAK)).^11–14^ Hence, understanding the role of DNAJC12 in monoamine synthesis may reveal novel strategies to reestablish monoamine levels in neurological disorders. A summary of the genotype–phenotype relationship is warranted in patients with DNAJC12 mutations, along with a literature review on DNAJC12 protein function, binding partners, and its role in monoamine synthesis.

### Monoamine Biosynthesis

Monoamines (include the catecholamines: dopamine, noradrenaline and adrenaline, and serotonin) fulfill multifunctional roles as neurotransmitters, trophic factors, hormones, and modulators of many physiological processes (Fig 1).^15–18^ The synthesis of catecholamines starts with the amino acid tyrosine which can be obtained from diet or produced in the liver from hydroxylation of phenylalanine by PAH. The catecholaminergic neurons and adrenal glands synthesize catecholamines according to cell-specific pathways, but with overlapping biochemistry using the same metabolic steps, enzymes, substrates and co-factors. In dopaminergic neurons, tyrosine is hydroxylated by TH to form dihydroxyphenylalanine (L-DOPA).^17,19^ Aromatic amino acid decarboxylase (AADC) decarboxylates L-DOPA to form dihydroxyphenethylamine (dopamine), which is the principal neurotransmitter of dopaminergic neurons. Adrenergic neurons that express the enzyme dopamine β-hydrolase (DH) convert dopamine into noradrenaline, whereas chromaffin cells in the adrenal medulla that express phenylethanolamine-N-methyl transferase (PNMT) convert noradrenaline to adrenaline.^17,19^ In contrast, serotonin synthesis involves the hydroxylation of an essential amino acid tryptophan to produce 5-hydroxytryptophan (5-HTP). This reaction is facilitated by TPH1 in the enterochromaffin cells (EC) of the gastrointestinal (GI) mucosa, pancreatic islets, mammary glands, and adipose tissue, and by TPH2 predominantely in the central nervous system (CNS). Subsequently, 5-HTP is decarboxylated by AADC to form serotonin. Notably, 90-95% of the total body serotonin is synthesized by EC.^16^ The synthesis of monoamines requires BH_4_ as a co-factor for PAH, TH, TPH1 and TPH2. BH_4_ is synthesized from guanosine triphosphate (GTP) with GTP cyclohydrolase 1 (GTPCH1, encoded by *GCH1*) as a rate-limiting enzyme, and deficiencies in its synthesis and recycling compromise the production of monoamines. In addition, DNAJC12 supports the synthesis of monoamines as a co-chaperone for PAH, TH, TPH1 and TPH2.^7,8^

**Fig 1.**
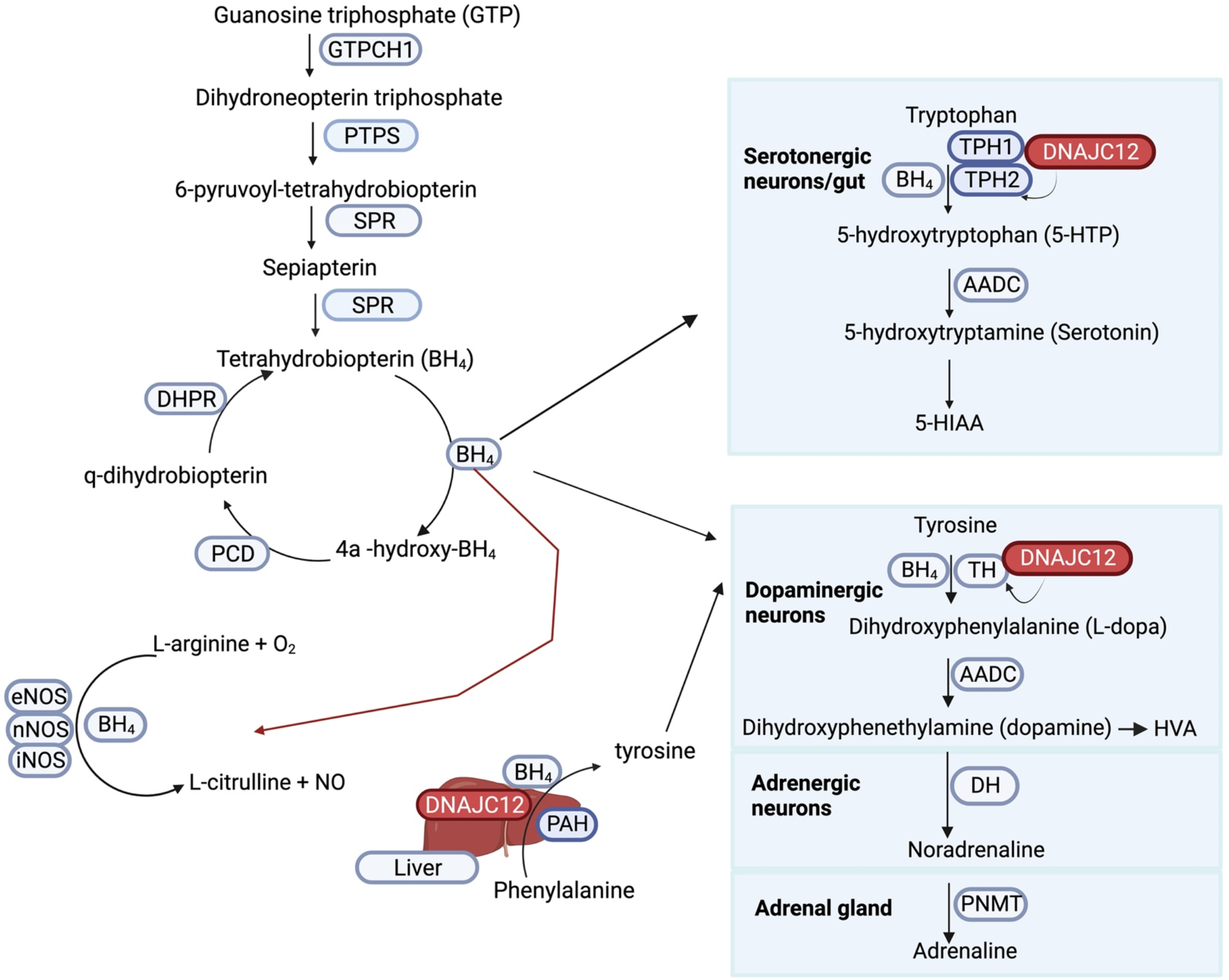
Monoamine biosynthesis. In a series of enzymatic reactions, GTP is converted into BH_4_, which acts as a co-factor for enzymes involved in the synthesis of monoamines and nitric oxide. DNAJC12 protein interacts with PAH, TH, TPH1 and TPH2, supporting its role in monoamines synthesis. Mutations in *DNAJC12*, and enzymes involved in BH_4_ synthesis and recycling, impair the synthesis of monoamines, resulting in overlapping neurological disorders.^7,8,19,20^ **Abbreviations:** GTPCH1: GTP cyclohydrolase 1; PTPS: 6-pyruvoyltetrahydropterin synthase; SPR: sepiapterin reductase; PCD: pterin-4a-carbinolamine dehydratase; DHPR: dihydropteridine reductase, eNOS: endothelial nitric oxide synthase; nNOS: neuronal nitric oxide synthase; iNOS: inducible nitric oxide synthase; PAH: phenylalanine hydroxylase; AADC: aromatic amino acid decarboxylase; TH: tyrosine hydroxylase; HVA: homovanillic acid; DH: dopamine β-hydrolase; PNMT: phenylethanolamine-N-methyl transferase; TPH1: tryptophan hydroxylase 1; TPH2: tryptophan hydroxylase 2; 5-HIAA: 5-hydroxindoleacetic acid. This figure was created with Bio-render.com.

### Disorders of monoamines

Disorders of monoamines due to defects in BH_4_ synthesis and recycling, enzymes involved in monoamine biosynthesis and catabolism, and monoamine transporter defects, lead to rare neurological disorders which manifest in childhood.^20^ Disorders of BH_4_ synthesis and recycling include autosomal dominant GTPCH1 deficiency caused by rare mutations in the *GCH1 gene*, autosomal recessive GTPCH1 deficiency, or may result from sepiapterin reductase (SR) deficiency, 6-pyruvoyltetrahydropterin synthase (PTPS) deficiency, dihydropteridine reductase (DHPR) deficiency or pterin-4a-carbinolamine dehydratase (PCD) deficiency. BH_4_ disorders are marked by a reduction in dopamine and serotonin, and consequently by a multitude of neurological complications, mostly co-occurring in the same patients, which include developmental delay, axial hypotonia, dystonia, parkinsonism, neuropsychiatric disturbances, cognitive disabilities and autonomic dysfunction.^21–23^ Of the BH_4_ disorders, PCD deficiency is the only one marked by the absence of severe neurological symptoms. Patients with BH_4_ disorders benefit immensely from treatment with neurotransmitter precursors (L-DOPA and 5-HTP) and/or BH_4_ therapy. However, the latter must be avoided in patients with defective BH_4_ recycling due to the potential accumulation of harmful BH_4_ metabolites.^20,24^ BH_4_ is also a co-factor for nitric oxide synthases and has been implicated in neuropathic and inflammatory pain. Pain is commonly reported in many neurodegenerative diseases and its management is based on clinical experience as there are limited treatment guidelines.^25^ *In-vivo*, axonal and peripheral injury elevates *GCH1* protein and activity, causing increased levels of BH_4_ and enhanced pain hypersensitivity. Such phenotypes can be ameliorated via knockout of *GCH1* or pharmacological inhibition of SR, which reduces the levels of BH_4_.^26,27^ In addition, it has been reported that certain polymorphisms in human *GCH1* protect against pain by reducing the levels of BH_4_, yet in a manner that maintains sufficient baseline activity of the enzyme such that the monoamine-associated neurological disorders are circumvented.^26–28^ Thus, further research to understand the BH_4_ pathway could unveil novel strategies to alleviate pain. Deficiency in the primary enzymes involved in monoamine biosynthesis such as TH and AADC, result in monoamine deficiency and neurological features that resemble BH_4_ disorders.^29,30^ These disorders, along with monoamine catabolism and transporter disorders, are reviewed in more detail by Brennenstuhl and colleagues.^20^ Genetic variants in the newly described molecular co-chaperone, DNAJC12, cause neurological features that resemble BH_4_ disorders and broaden the spectrum of monoamine disorders ^20^, as elaborated below.

### DNAJC12 structure and binding partners

The *DNAJC12* gene is located on chromosome 10q21.3. It has five exons and can be alternatively spliced to generate two transcripts.^31^ The longer *DNAJC12* transcript encodes the full-length protein of 198 amino acids, while a shorter transcript encodes a protein of 107 amino acids.^31^ DNAJC12 protein is expressed predominately in the cytoplasm; it contains a J-domain and a highly conserved C-terminus without other known functional domains (Fig 2A).^31^ It is prominently expressed in the brain, adrenal glands and liver (https://www.proteinatlas.org/ENSG00000108176-DNAJC12/tissue#rna_expression). While the role of DNAJC12 has yet to be fully elucidated, it is widely regarded as a co-chaperone for AAAHs such as PAH, TH, TPH1 and TPH2.^2,7^ DNAJC12 interacts with PAH *in-vitro* and *in-vivo,* and as a corollary, PAH protein and activity are reduced in the fibroblasts of patients with *DNAJC12* mutations ^7,32^ while dopamine and serotonin are reduced in their cerebrospinal fluid, consistent with the role of DNAJC12 as a co-chaperone for TH and TPH2.^7,33^ Notably, DNAJC12 is a binding partner of BiP/GRP78, a critical ER HSP-70 chaperone that is upregulated with ER stress.^31^ The ER transcription factor AIbZIP also increases DNAJC12 protein expression during ER stress, exerting its effects on the *DNAJC12* promoter.^31^ As HSC-70 interacts with TH, AADC and the vesicular monoamine transporter (VMAT2), DNAJC12 appears to be indirectly involved in dopamine synthesis and packaging ^34–36^, however further work is necessary to validate this connection.

**Fig 2.**
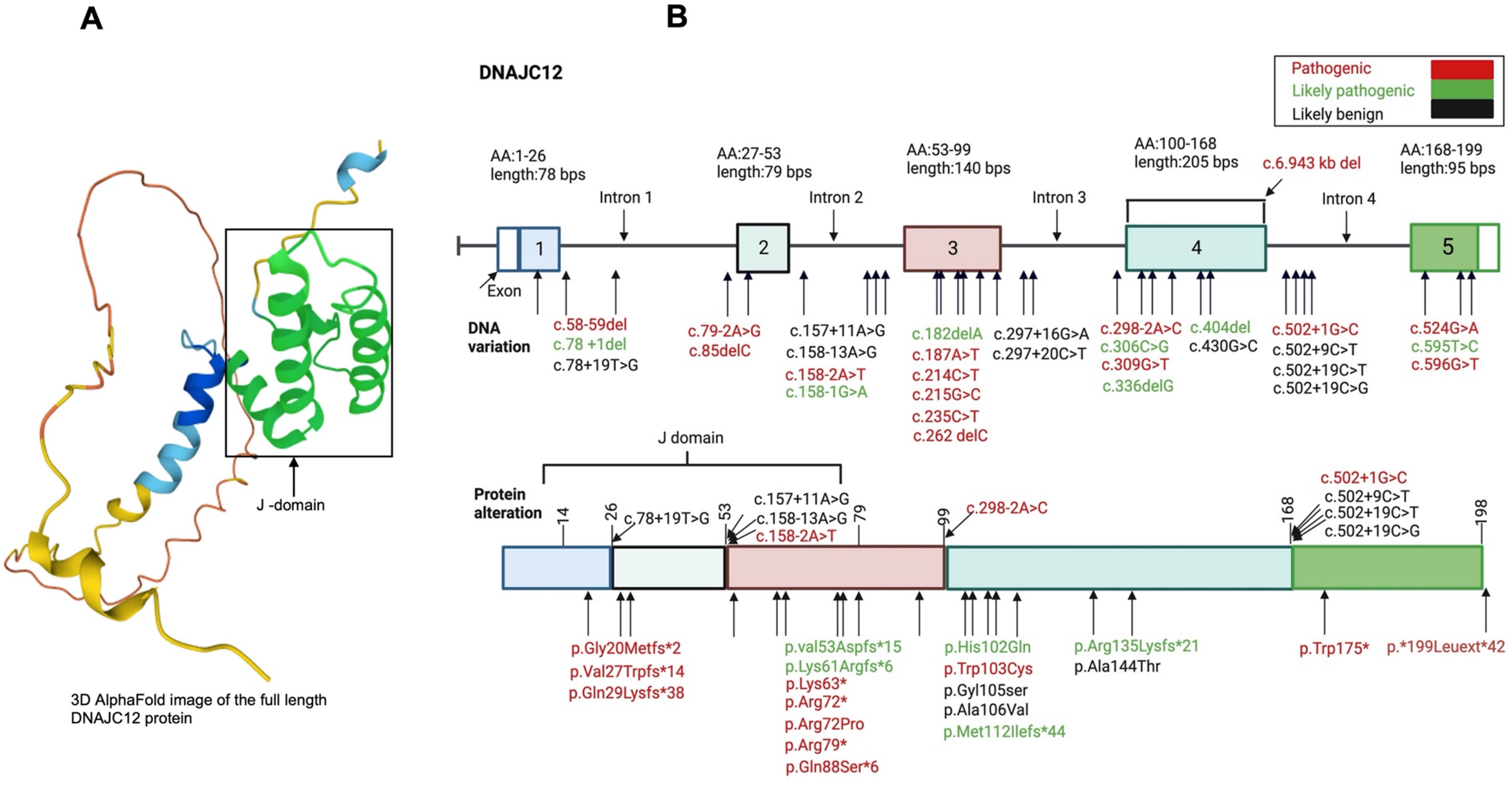
Genetic variants in *DNAJC12*. **A)** 3D image of DNAJC12 protein from AlphaFold. **B)** Variants in *DNAJC12* gene (top image), and the corresponding protein alteration (bottom image). Variants in *DNAJC12* gene and protein were identified in published articles, and within Clinvar, GnomAD and Uniprot databases. They were classified as pathogenic, likely pathogenic or likely benign according to the guidelines of the American College of Medical Genetics and Genomics, and Association of Molecular Pathology.^37^ This figure was created with Bio-render.com.

### Genotype and phenotype relationships in patients with *DNAJC12* mutations

#### Literature search and selection criteria

A literature search was performed using PubMed and Google Scholar with the following keywords: hyperphenylalaninemia or phenylketonuria, DNAJC12 or heat shock protein 40 (HSP40). This search yielded 216 articles which included a total of 639 patients with HPA (including patients with non-BH_4_ or BH_4_ HPA). After four patients were excluded with heterozygous mutations in PAH, 52 patients (8.1%) with HPA were found to have biallelic or heterozygous mutations in *DNAJC12*. Clinical data was ultimately extracted from 15 articles published between 2017 and 2023. *DNAJC12* variants are classified as pathogenic, likely pathogenic and likely benign according to the guidelines of the American College of Medical Genetics and Genomics and Association of Molecular Pathology ^37^, and were also identified within Clinvar, GnomAD and Uniprot databases (Fig 2B).

### Overview of patients with *DNAJC12* mutations

Biallelic pathogenic variants in *DNAJC12* were first identified via whole exome sequencing in six affected children from four unrelated consanguineous families from Morocco, Turkey, and Saudi Arabia.^7^ Five of the six patients were investigated because of a neonatal/pediatric movement disorder resembling a BH_4_ deficiency syndrome with HPA, infantile dystonia and progressive neurodevelopmental delay. High-throughput sequencing excluded mutations in *PAH*, whereas deficiencies in BH_4_ were ruled out by assessing urinary pterin metabolites and DHPR activity in dried blood spots. One of the six individuals was immediately treated with a combination of neurotransmitter precursors (L-DOPA and 5-HTP), BH_4_ and folinic acid due to prior history of HPA within their family, and they were spared from neurological deficits.^7^ Such findings suggest that much of the neurodevelopmental phenotype attributed to *DNAJC12* mutations can be prophylactically managed via early diagnosis and timely treatment. A second independent study in Canadian and Italian families used comparative whole exome sequencing to successfully identify biallelic *DNAJC12* mutations as a cause of young-onset parkinsonism.^8^ It appeared that HPA was only apparent in one of three affected individuals. However, prominent features in all patients were mild intellectual disability and L-DOPA-responsive nonprogressive parkinsonism with dyskinesia, which were not associated with dopaminergic deficits by presynaptic nigrostriatal imaging.^8^ Thus, these patients might constitute cases of “scans without evidence of dopaminergic deficits” an enigmatic clinical dilemma that is also evident in patients with Parkinson’s disease (PD) and levodopa-responsive dystonia.^38–40^ With the recognition that recessively inherited *DNAJC12* loss-of-function mutations may cause HPA, this gene has recently been included in newborn genetic testing.^41–44^ Recognizing this bias and based on patients identified with *DNAJC12* mutations between 2017–2023 (Fig 3A to B), the most predominant features in the first decade include developmental delay, attention deficit hyperactivity disorder (ADHD), speech delay, intellectual disability, autism, axial hypotonia and dystonia. In the second decade, the phenotype is marked by prominent parkinsonism, dystonia, axial hypotonia, with/without intellectual disability, and to a lesser extent, psychiatric features, ADHD or other cognitive deficits. Clinical data from the third decade is scant, given a lack of genetic testing in this age group, but the symptomology includes young-onset parkinsonism, cognitive deficits and psychiatric features.

**Fig 3.**
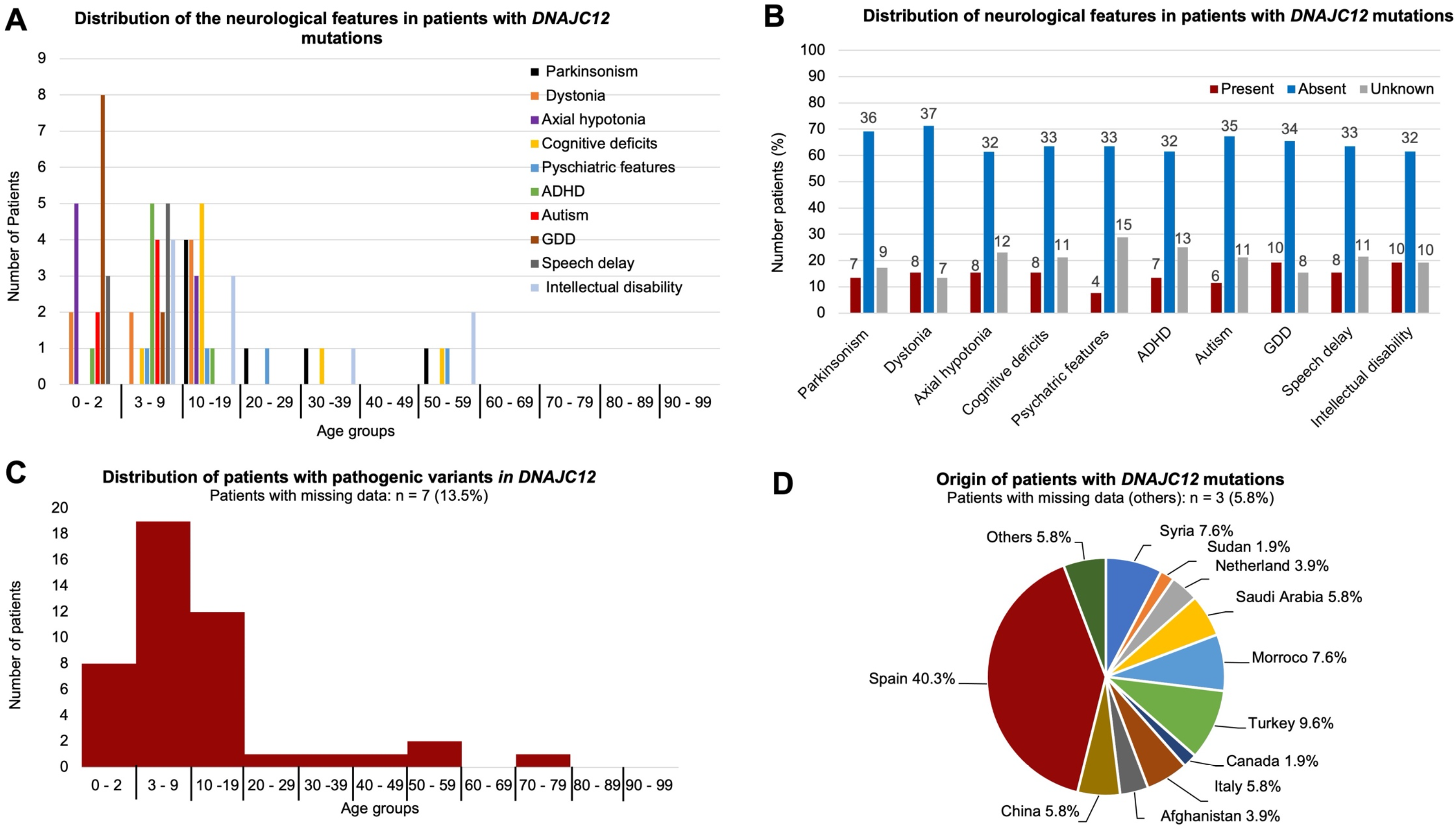
Distribution of patients with *DNAJC12* mutations. Clinical data was extracted from 15 articles published between 2017 to 2023, with a total of 52 patients. Only the most common neurological features are depicted in this figure. **A)** Number of patients with neurological features by age group. **B)** Number of patients with, without or of unknown status for a specific neurological feature. **C)** Distribution of patients with *DNAJC12* mutations by age group. **D)** Origin of patients with *DNAJC12* mutations. Note that “others” on the pie chart refers to the percentage of patients with undefined origin. **Abbreviations:** ADHD: attention deficit hyperactivity disorder; GDD: global developmental delay.

### Genetics variants and clinical features of patients with *DNAJC12* mutations

A total of 52 patients was identified with biallelic or heterozygous *DNAJC12* mutations originating from 11 countries (missing data: 3 patients) located in Europe, Asia, the Middle East, and North Africa (Fig 3D). Seventy-five percent of patients (39 patients) were homozygous and 25% (13 patients) were compound heterozygotes. The types of *DNAJC12* mutations included nonsense (34.6%, 18 patients), splice site (26.9%, 14 patients), frameshift (11.5%, 6 patients), missense variants (1.9%, 1 patient) and others, which refer to compound heterozygous variants with two different types of mutations (25%, 13 patients) (Fig S1). Neurological features vary markedly among patients with *DNAJC12* mutations, even within siblings with the same genotype, and the age of onset ranged from 22 months to 59 years. Detailed clinical data is provided in Table S1. In brief, patients with *DNAJC12* mutations had movement disorders exemplified by axial hypotonia (15.4%, 8 patients), dystonia (15.4%, 8 patients) and parkinsonism (13.5%, 7 patients), which co-exist in some patients.^7,8,44–46^ Recessively inherited, biallelic *DNAJC12* variants are a rare cause of young-onset L-DOPA-responsive parkinsonism; ^8^ although heterozygous variants neither appear to be a risk factor nor a genome-wide associated locus for idiopathic PD.^47–49^ Other neurological features include global developmental delay (19.2%, 10 patients), intellectual disability (19.2%, 10 patients), cognitive deficits (15.4%, 8 patients), speech delays (15.4%, 8 patients), ADHD (13.5%, 7 patients), autism (11.5%, 6 patients) and other psychiatric features (7.7%, 4 patients).^42,43,50^ Many of these features co-occur in the same patients and are developmental or associated with age. Of note, 40.4% (21 patients) of patients with *DNAJC12* mutations (ranging from 6 – 40 years old) carried a homozygous (13 patients) or heterozygous (8 patients) p. Trp175* variant, which was only reported in patients with a Spanish origin.^51,52^ Increased cases of DNAJC12-induced HPA recognized in the Spanish population reflect this country’s comprehensive implementation of *DNAJC12* sequencing in newborn genetic screening.^51^ Of the patients with homozygous p. Trp175* variants in *DNAJC12*, 2 had unknown neurological status, 10 were neurologically asymptomatic, and only 1 patient was reported with neurological features, which included ADHD, autism and intellectual disability. Conversely, patients with a heterozygous p. Trp175* variant included 2 that were asymptomatic, 3 symptomatic and 3 with an unknown neurological status. These findings suggest that specific genetic or environmental factors govern the penetrance of p. Trp175* in the Spanish population. Of note, p. Trp175* is a nonsense variant that results in a premature stop codon in the last exon of the *DNAJC12* gene, which is predicted to result in a protein that is 23 amino acids shorter at the C-terminus. Although it hasn’t been examined in homozygous p. Trp175* patients, DNAJC12 protein is markedly reduced in the fibroblasts of heterozygous patients compared to matched controls.^51^ Notably, 42.3% of individuals with DNAJC12 mutations (22 patients) were reported to be neurologically asymptomatic. Some individuals described as asymptomatic refused to perform neuropyschometric tests, and in some, it was unclear whether more informative exams might have identified more subtle neurological features. Therefore, some phenotypes may have been missed in seemingly asymptomatic individuals with *DNAJC12* mutations.

### Diagnosis and treatment in patients with *DNAJC12* mutations

*DNAJC12* mutations are mostly found when newborns with HPA undergo whole exome sequencing or when individual patients are investigated for developmental delay with motor complications.^7,8,33^ *DNAJC12* mutations elevate the levels of Phe in the blood and result in dopamine and serotonin deficiency. DNAJC12 patients would continue to experience a progressive increase in Phe levels without intervention since they cannot correctly metabolize this amino acid, which has severe neurological consequences.^9^ As a result, the standard treatment in patients with *DNAJC12* mutations is a Phe-restricted diet to maintain low levels of Phe in children, adolescents and adults.^9^ The literature is variable regarding the blood Phe concentration required to start treatment. There is a consensus that patients with untreated blood Phe concentration of >600 μmol/L should commence treatment while patients with <360 remain untreated. Unfortunately, it is unclear whether patients with untreated blood Phe concentration >360 should be treated.^53,54^ Only one study reported that patients with untreated blood Phe concentration >360 have similar neuropsychological scores compared to control individuals, except in their executive function.^55^ A Phe-restricted diet is often coupled with oral dopamine and serotonin precursors (L-DOPA and 5-HTP, respectively) to compensate for neurotransmitter deficiency and may include other supplements, including sapropterin dihydrochloride (a synthetic BH_4_), folinic acid, selegiline, entacapone and pramipexole. Such treatment has ameliorated neurological features in 12 of 18 patients, with those in their first three months of life responding best with almost complete restoration of neurological function.^7,33,42,50,56,57^ Thirteen percent of the patients (7 patients) had BH_4_ treatment with or without a Phe-restricted diet, and 4 responded positively, including improved cognition and motor function, and 3 were allowed to return to their unrestricted diet.^7,33^ The beneficial effects of BH_4_ may be attributed to its role as a co-factor for AAAHs, enhancing the stability of PAH, TH and TPH2.^58^ Eight percent of the patients (4 patients) were treated with oral L-DOPA, possibly combined with a Phe-restricted diet.^8^ Of these, 3 had L-DOPA-responsive parkinsonism but developed dyskinesia. Nevertheless, in 9.6 % (5 patients), the treatment status is unknown ^52^, and 32.7% (17 patients) have yet to commence any treatment.^51,59^ While all untreated individuals had mild HPA, most were otherwise asymptomatic.

### Proposed pathological mechanisms in DNAJC12 patients

The pathogenesis of neurological problems in patients with *DNAJC12* mutation is not fully understood. A Phe-restricted diet alleviates many neurological features, complementary to findings for phenylketonuria, a disorder of Phe metabolism caused by PAH deficiency, therefore suggesting that HPA is similarly toxic.^33,50,51^ Multiple mechanisms underlying those effects have been proposed, from deficits in lipid metabolism, cholesterol synthesis and oligodendrocyte myelination, to impaired glucose metabolism and large amino acid transport, to monoamine deficiency. Nevertheless, the relative contribution of *DNAJC12* loss-of-function to patients’ phenotypes has yet to be elucidated.

### White matter lesions

White matter disruption is strongly associated with HPA, and to our knowledge, has yet to be investigated in patients with *DNAJC12* mutations.^33^ The precise mechanisms underlying white matter abnormalities are not well understood. Nonetheless, a leading hypothesis is that HPA impairs myelination in the brain.^9,60,61^ Congruent with this hypothesis, HPA changes the phenotype of oligodendrocytes from myelinating to non-myelinating in mice with biallelic *PAH* mutations.^9,62^ Such findings are consistent with *in-vitro* experiments showing that HPA reduces myelin basic protein in cerebellar organotypic slices.^63^ However, other *in-vitro* studies claim oligodendrocytes are capable of synthesizing myelin sheath under HPA, suggesting other mechanisms are involved in demyelination.^64^ A portion of the myelin lipid is made up of cholesterol, and an alteration in cholesterol synthesis could impair the formation of myelin sheath; hence, hypomyelination. In PAH-deficient mice, HPA inhibits the activity of 3-hydroxyl-3-methglutaryl coenzyme A reductase (HMGR), a rate-limiting enzyme in the synthesis of cholesterol in the forebrain. There was also a reduction in HMGR activity and cholesterol levels in the forebrain of PKU mice.^65^

### Deficiency in monoamines

Deficiency in central dopamine and serotonin is a core feature of patients with *DNAJC12* mutations, and this is congruent with the role of DNAJC12 as a co-chaperone for monoamines. ^7,8,33^ HPA may also inhibit the influx of large neural amino acids (LNAAs) into the brain through Phe-mediated competition.^9,66^ These LNAAs include tyrosine and tryptophan which are essential for the synthesis of dopamine and serotonin, respectively. The synthesis of the latter monoamines could also be instigated by Phe-induced competitive inhibition of TH and TPH2 activities.^67^ Catecholamines such as noradrenaline and adrenaline are downstream of dopamine in the catecholamine synthesis pathway, and may also be affected by DNAJC12 deficiency. However, these neurotransmitters/hormones have yet to be examined in patients with *DNAJC12* mutations. Dopamine is essential for movement, attention, addiction, reward, cognition, arousal, wakefulness, memory formation and consolidation.^17,68–70^ These functions are under the control of three large dopaminergic neural circuits in the CNS; namely, the nigrostriatal pathway, the mesocortical pathway and the mesolimbic pathway.^17^ The nigrostriatal pathway consists of midbrain dopaminergic neurons in the *substantia nigra pars compacta* (SNpc), which send their axons to the caudate and putamen, to primarily control movement initiation. The mesocortical pathway consists of midbrain dopaminergic neurons in the adjacent ventral tegmental area (VTA), which project to the prefrontal cortex (PFC) that is pivotal for executive skills, attention, planning and decision-making. The mesolimbic pathway also originates in the VTA, whereby dopaminergic neurons send their projections to the hippocampus, amygdala and nucleus accumbens, with a role in processing and formation of emotion, learning, working memory and long-term memory. Hence, dysfunction in dopaminergic neural circuits may have multiple roles in the neurological problems experienced by DNAJC12 patients, such as parkinsonism, dystonia, ADHD, intellectual disability and cognition. It is important to note that catecholamines have an essential role in the periphery which may also be compromised by *DNAJC12* mutations. Secreted into the blood as hormones in the fight or flight response, the catecholamines increase pupil dilation, heart rate, blood sugar levels and blood flow to muscles. Also, they have direct effects on peripheral organs including the heart, blood vessels, kidney, liver, pancreas, stomach, brown adipose tissue and the immune system.^15,18,71,72^

Serotonin in the CNS is severely reduced in patients with *DNAJC12* mutations, and this neurotransmitter is integral in neurite outgrowth, synaptogenesis and neurogenesis.^73^ Serotonergic innervation originates predominately in the raphe nuclei in the brainstem, and is widely distributed throughout the CNS.^74^ Impaired serotonin is well documented in neurodevelopmental disorders such as autism and in neuropsychiatric and mood disorders, including anxiety, apathy and depression, all of which are among the plethora of neurological features documented in patients with *DNAJC12* mutations.^73–76^ Serotonin is involved in GI motility and inflammation, and also functions in muscle constriction in the lung and uterus. It affects blood vessel constriction and dilation, platelet aggregation and nociceptive pain, all of which may be impacted by *DNAJC12* mutations ^77,78^. The diverse effects of serotonin in the periphery and brain, make it a potential regulator of the gut-brain axis. Considering the pleotrophic functions of monoamines, a conditional cre/loxP Dnajc12 knockout model would be a singular, but comprehensive approach to study catecholamine communication between the periphery, basal ganglia and cortex.

## Discussion

*DNAJC12* mutations were discovered in 2017 as a new cause of HPA with neurodevelopmental and motor disorders.^7,8^ Newborn screening for *DNAJC12* mutations can now identify cases of mild HPA, unexplained by mutations in PAH and BH_4_, and can ensure earlier treatment. Most disease-causing mutations appear to be recessively inherited and are a loss-of-function. However, variants such as p.Trp175* may retain partial function and have reduced penetrance. All patients with *DNAJC12* mutations, irrespective of their neurological symptoms, should start treatment to reduce Phe if diagnosed with HPA. However, maintaining an adequate protein diet is important in developing children and is challenging.^9^ Pegvaliase, a bacterial-derived phenylalanine ammonia lyase (PAL), conjugated to polyethylene glycol (PEG) to reduce immunogenicity ^54,79^, was recently approved by Food and Drug Administration for the treatment of PKU, and it provides a new approach for treatment in patients with *DNAJC12* mutations.^79,80^ Pegvaliase converts Phe to trans-cinnamic acid and ammonia, and is highly effective at reducing blood Phe levels such that patients can return to an unrestricted diet. However, many *DNAJC12* patients may be within the normal range of Phe concentration (<360 μmol/L) and may be treated with neurotransmitter precursors alone or synthetic BH_4_, L-DOPA, folinic acid and selegiline, and may remain on an unrestricted Phe diet which is preferable in early development. Nevertheless, HPA should be routinely monitored given the neurotoxicity of elevated Phe in blood and prospective evaluation is warranted. Untreated, *DNAJC12* mutations are a rare cause of infantile dystonia and young-onset parkinsonism that can be effectively managed with BH_4_ or L-DOPA/5-HT replacement therapy.^7,8,33,81^ Adults with biallelic *DNAJC12* mutations may manifest with young-onset Parkinson’s disease but not have HPA. Curiously, dystonia and parkinsonism co-exist in several neurological disorders, and they can be an alternate manifestation of the same underlying cause (a *forme fruste*) and an adverse response to therapy (drug-induced dyskinesia). For example, heterozygous mutations in *GCH1,* encoding GTPCH1, the rate-limiting enzyme in the synthesis of BH_4_, may either manifest with postural dystonia in childhood which later generalizes, or as late-onset PD. Either syndrome may present, and even within different members of the same family.^82,83^ *GCH1* is also a genome-wide associated locus for sporadic, idiopathic late-onset PD. Although clearly related to catecholamine metabolism, the pathological mechanism for these alternate phenotypic presentations remains enigmatic.^12,38,81^

Biochemically, *DNAJC12* is a co-chaperone for monoamine synthesis, yet its role in the intermediary metabolism of these neurotransmitters and hormones has yet to be elucidated.^8^ However, temporal and tissue-specific models of *DNAJC12* knock out may further our understanding of the pleiotrophic functions of monoamines in the brain and periphery, and could reveal novel strategies to normalize levels in pathological states. Conditional models could help decipher the ancient, integrated systems biology that monoamines modify, from times of acute and chronic stress, to excess in many psychiatric conditions (addiction, ADHD, autism and mood disorders), to deficits in neurologic and neurodegenerative disorders (dystonia, dementia and parkinsonism). For example, dopamine and other catecholamines, such as noradrenaline, modify PFC function, including attention, arousal and executive function.^84^ Catecholamines such as dopamine may also have immunomodulatory effects as immune cells express dopamine receptors, although studies in the periphery are in their infancy.^85^

## Conclusion

This review highlights the role of *DNAJC12* variants in monoamine disorders and mild HPA; thus, validating its inclusion in newborn genetic screening. HPA is associated with neurotoxicity and neurodevelopmental disorders that can be alleviated by early diagnosis and treatment. Nevertheless, limited screening in the adult population means the full spectrum of clinical symptoms has yet to be fully appreciated. Dopamine deficiency is a prominent feature in patients with *DNAJC12* mutations that coincides with motor deficits such as dystonia and parkinsonism, and which is implicated in PFC dysfunction and impaired executive function.^7–9^ Serotonin is also drastically reduced in DNAJC12 patients and relevant to mood disorders, including anxiety, apathy and depression, which are a common neuropsychiatric problems manifest in patients with idiopathic Lewy body disorders.^86^ By documenting *DNAJC12* mutations in patients and related genotype-phenotype associations we highlight the potential for earlier diagnosis and intervention. Also, we highlight the utility of *in-vivo* models of *DNAJC12* to investigate the role of central and peripheral monoamines, their systems biology, and suggest knowledge of their integrated mechanisms of action is needed to inform the development of novel therapeutic strategies.

## Data Availability

All data produced in the present work are contained in the manuscript.

## Supplementary figure legends

**Fig S1.**
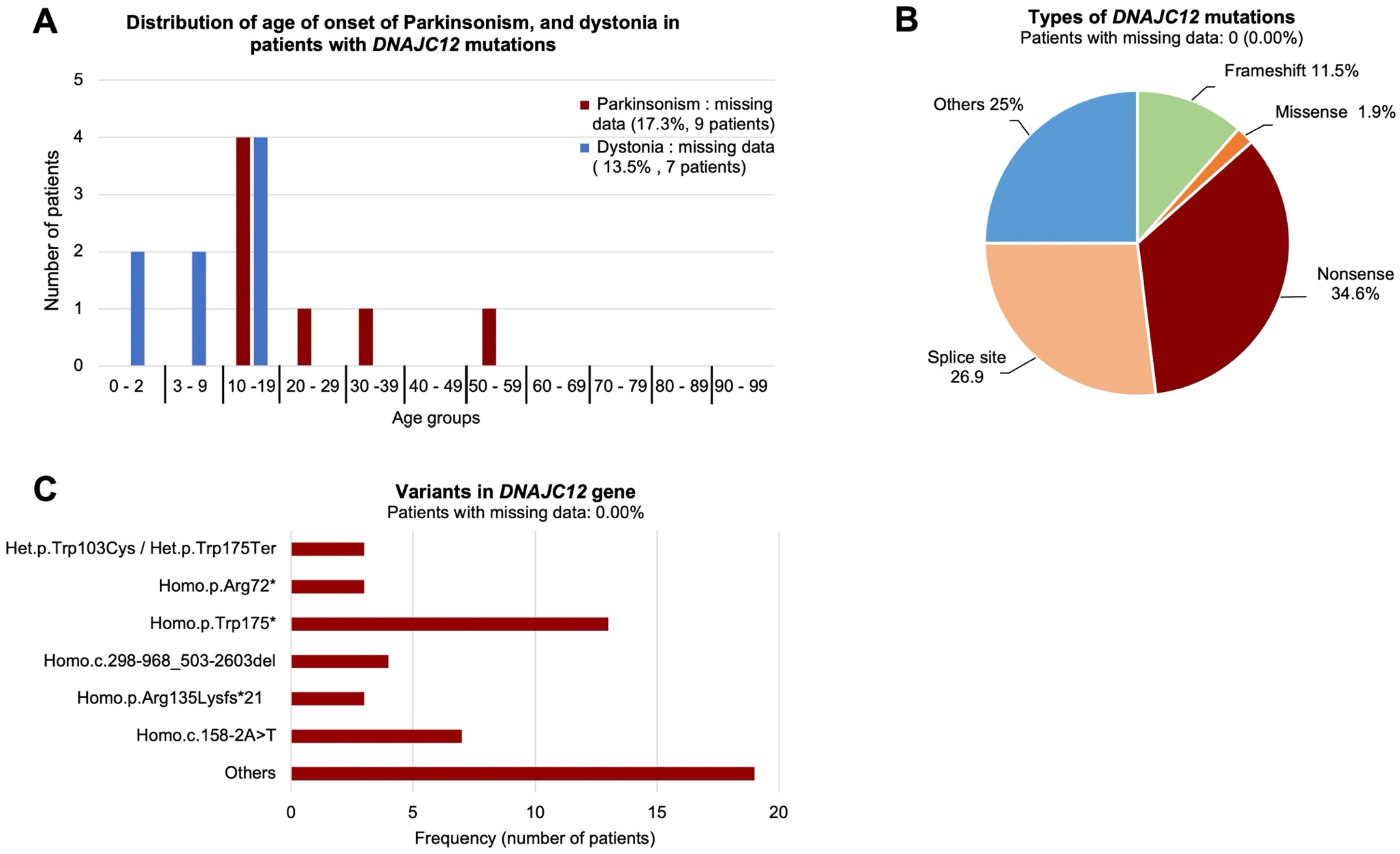
Distribution of patients with parkinsonism and dystonia, and types and individual variants in *DNAJC12*. **A)** Number of patients with parkinsonism and dystonia per age group**. B)** The types of mutations in *DNAJC12*. Note that "others" on the pie chart refers to compound heterozygous variants with two different types of mutations. **C)** Variants in *DNAJC12* gene/protein. Only the most frequent variants are presented individually, and the remaining variants are shown under "others".

## Supplementary tables

**Table S1. Detail clinical data of all the individuals identified with DNAJC12 mutations.** Clinical data was extracted from 15 articles published between 2017 to 2023, with a total of 52 patients. Their demographics, variants in *DNAJC12*, symptomology and treatment paradigms are detailed.

## Author contributions

IBD contributed to conception and design of the review, acquisition and analysis of data, and drafted a significant portion of the manuscript or figures. MJF contributed to conception and design of the review, and drafted a significant portion of the manuscript or figures. JF and MB drafted a significant portion of the manuscript.

---

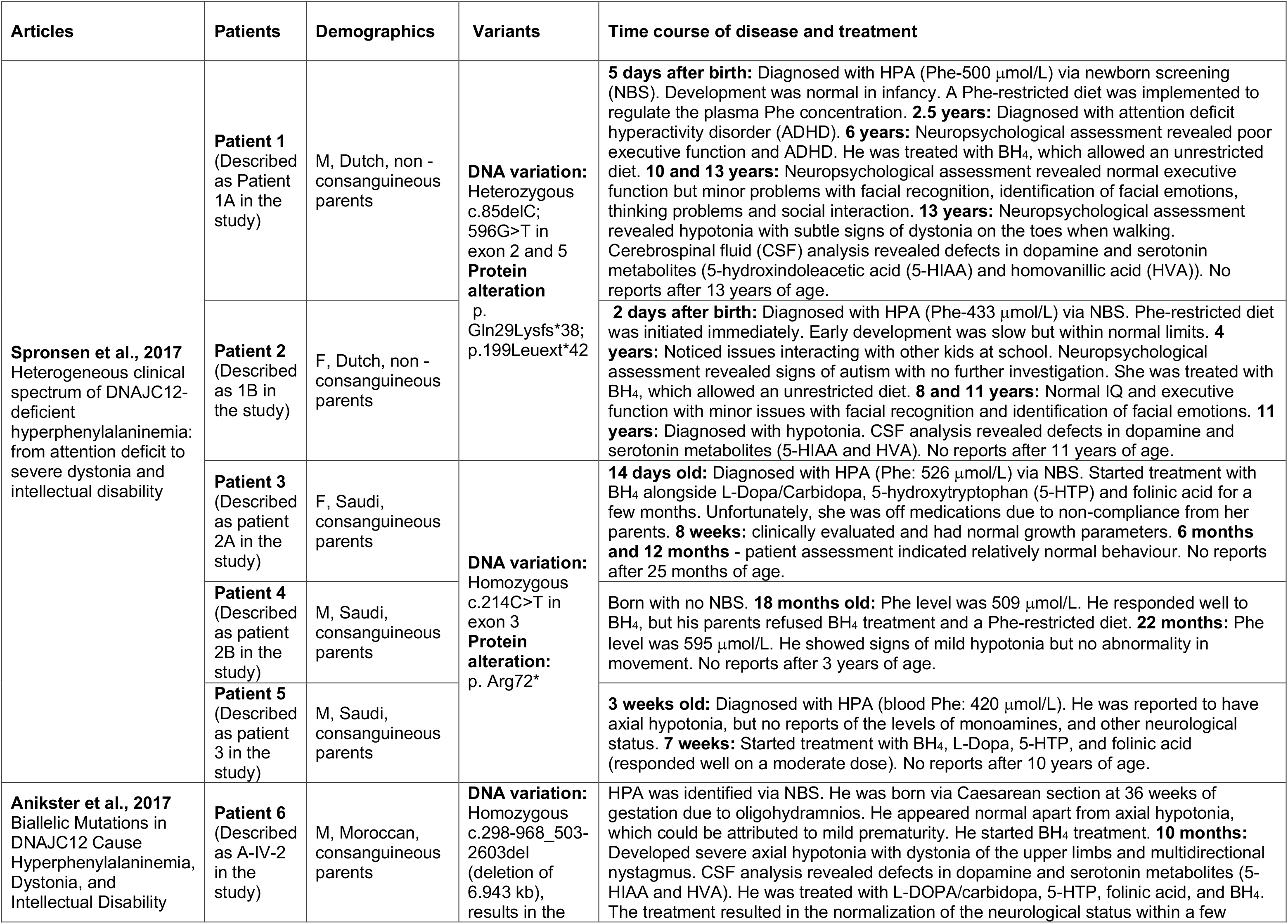

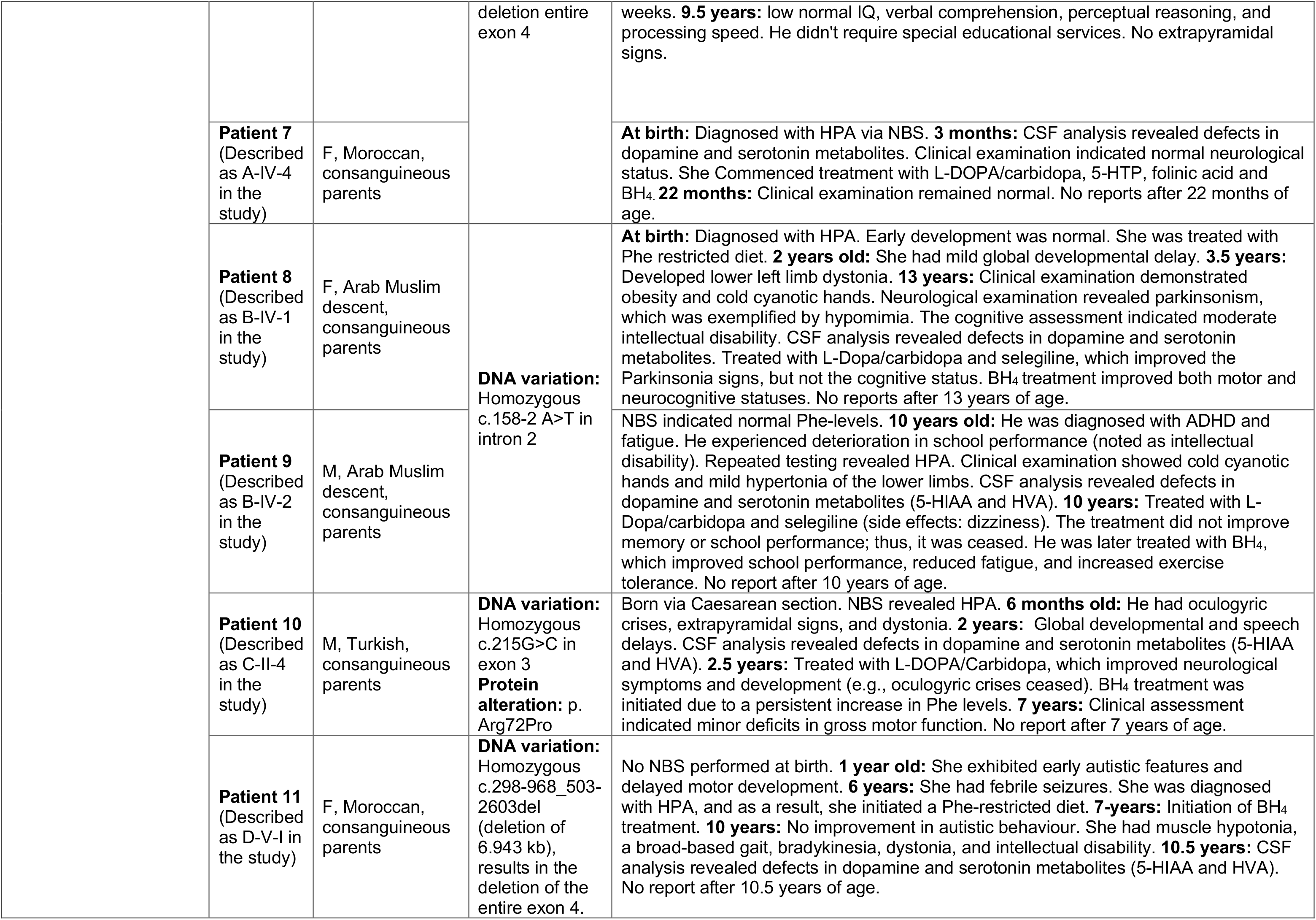

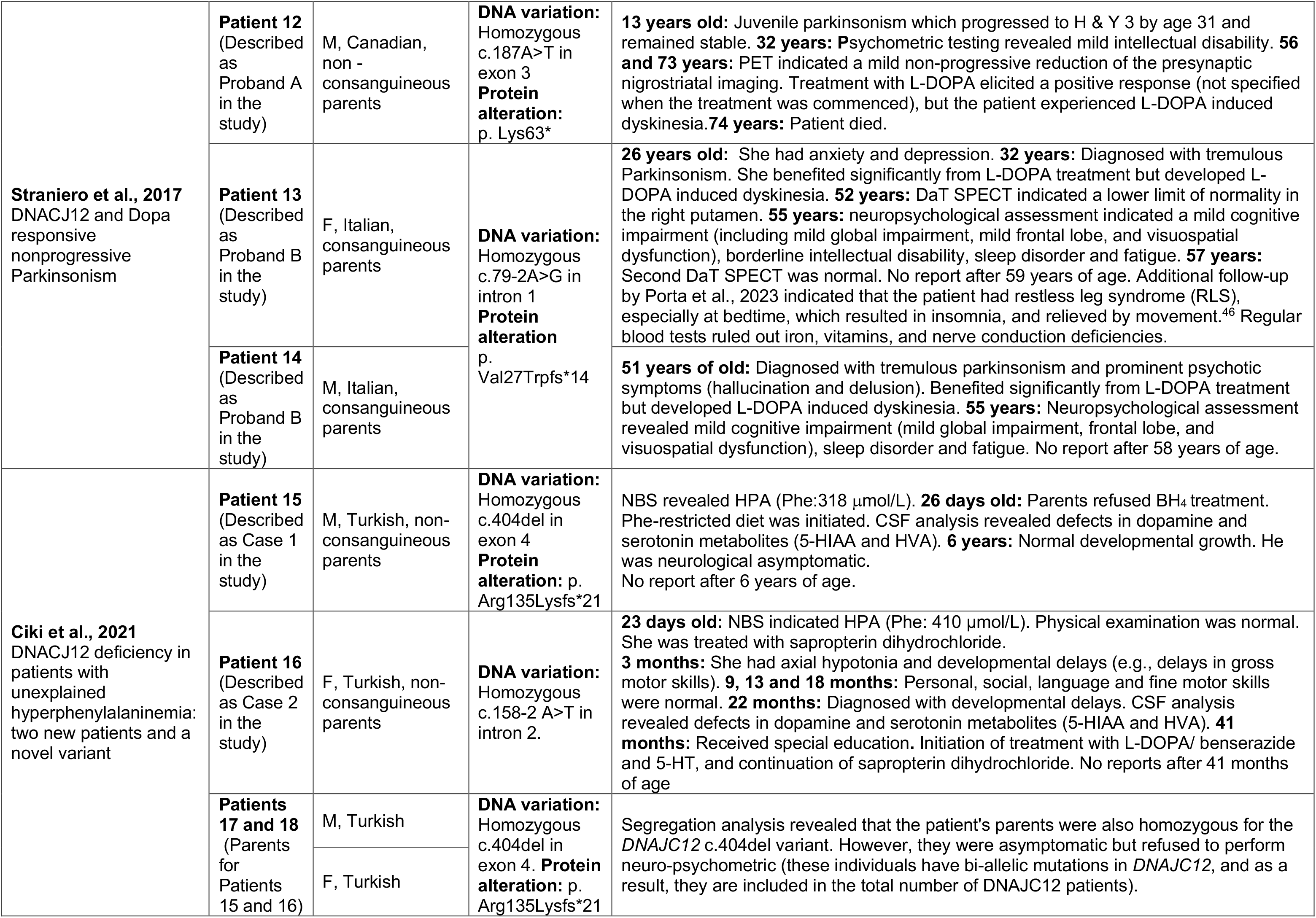

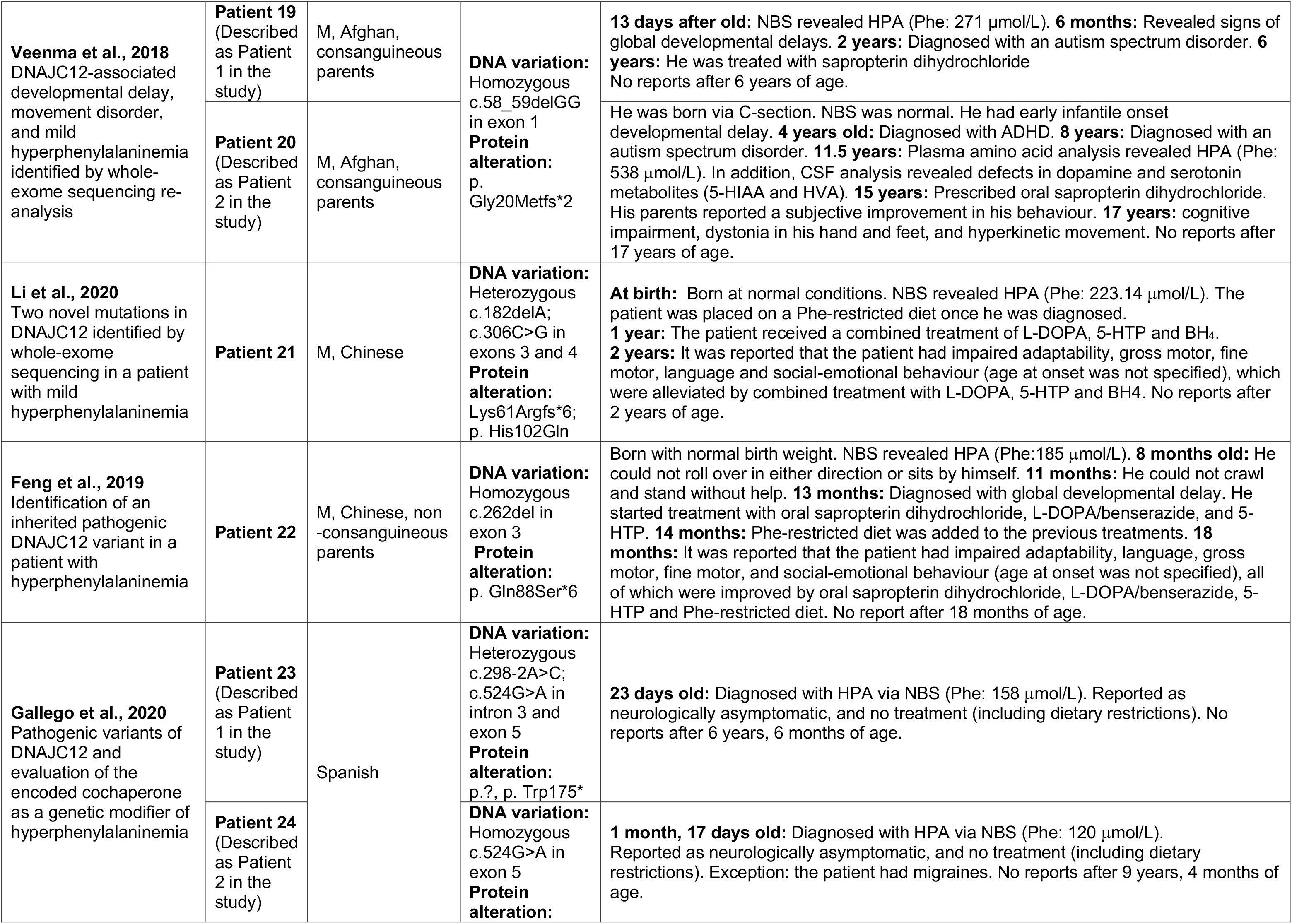

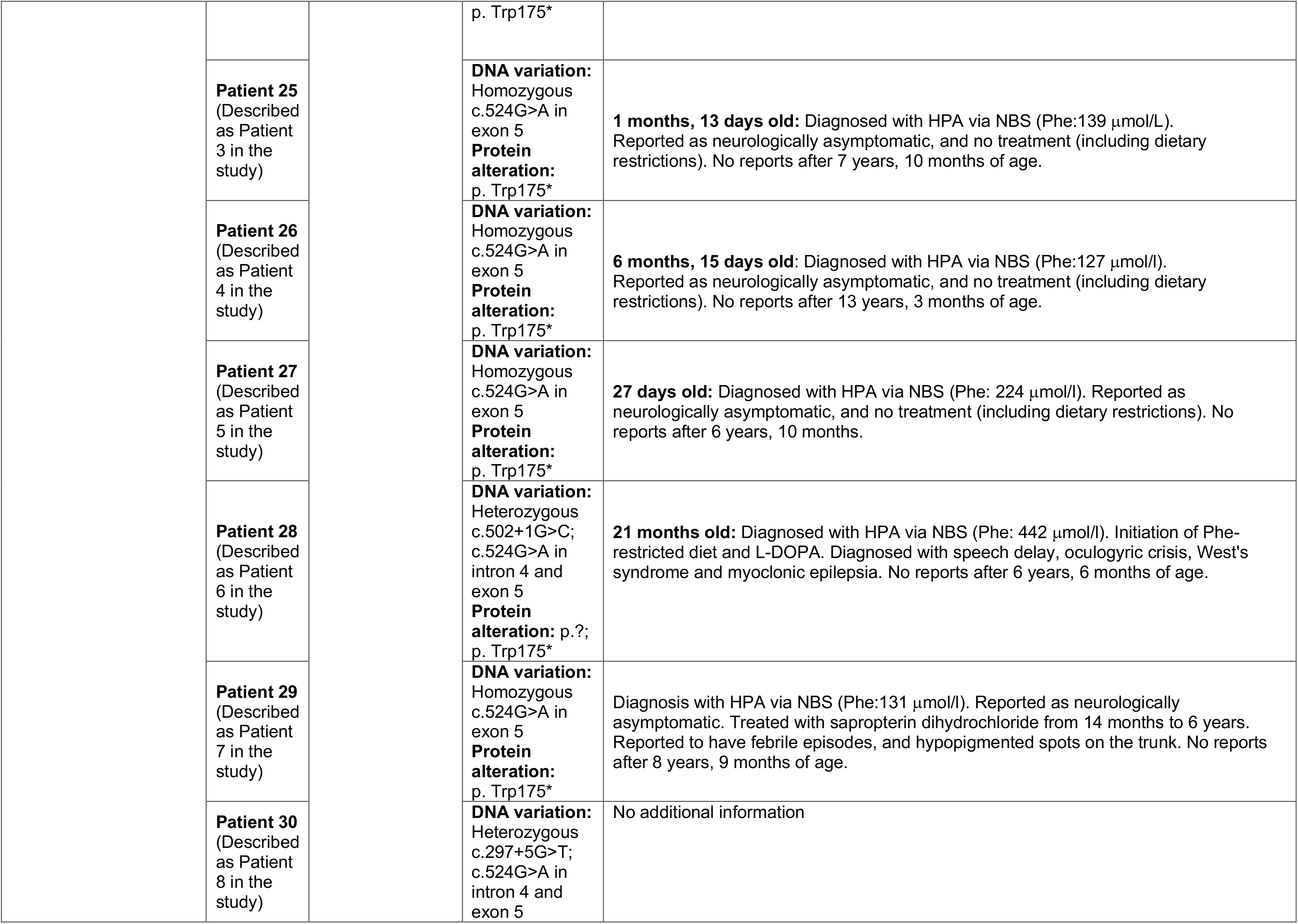

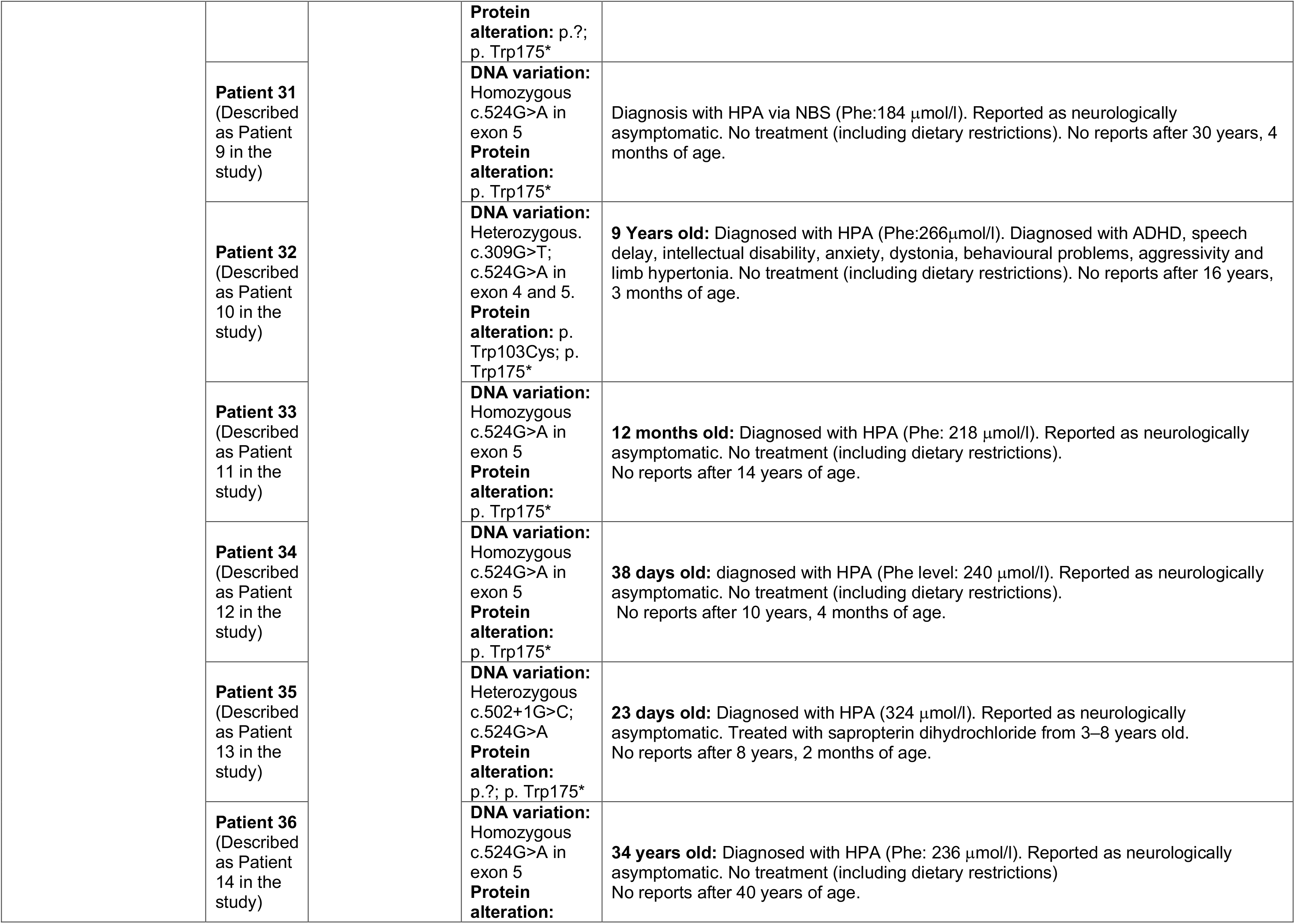

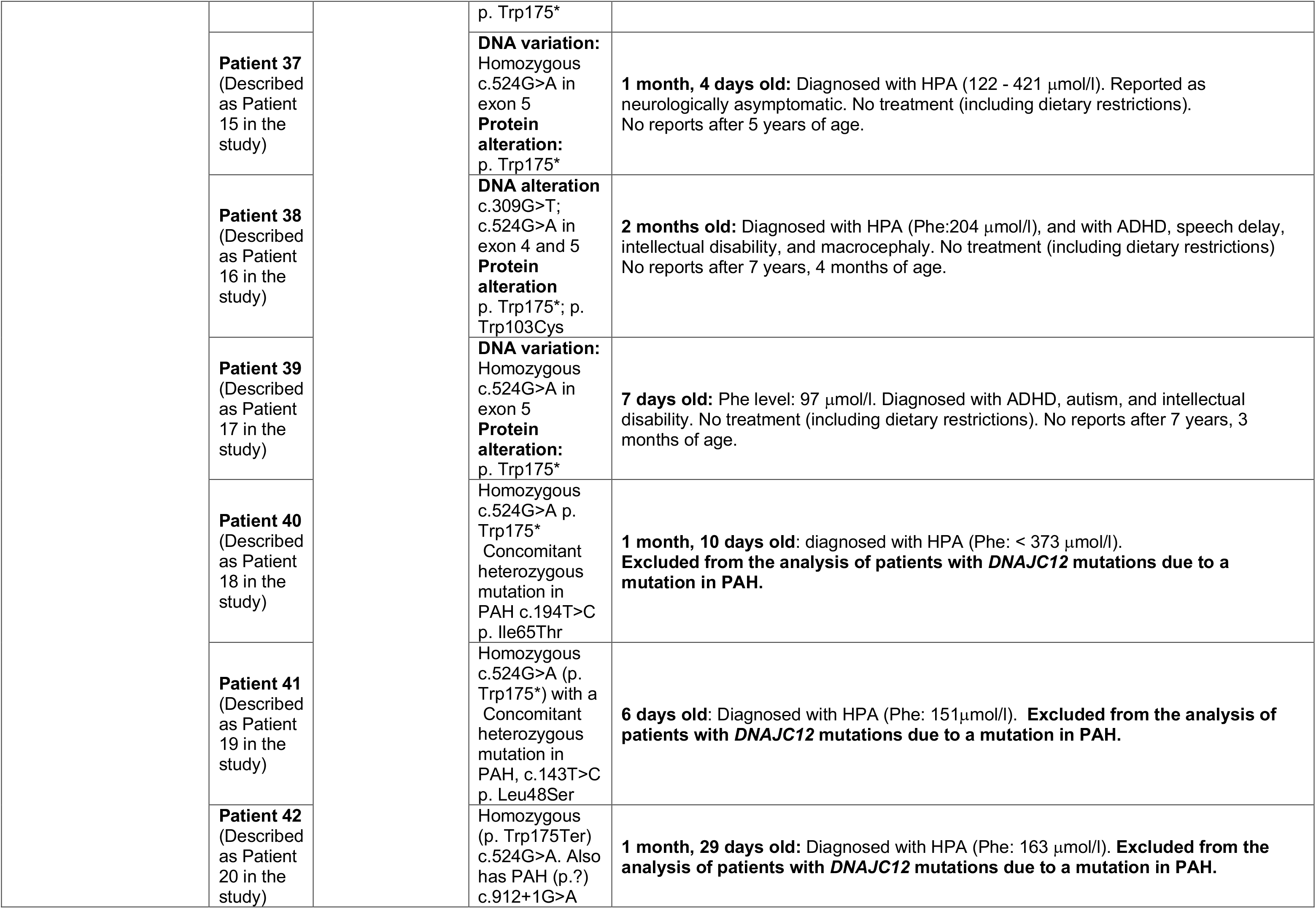

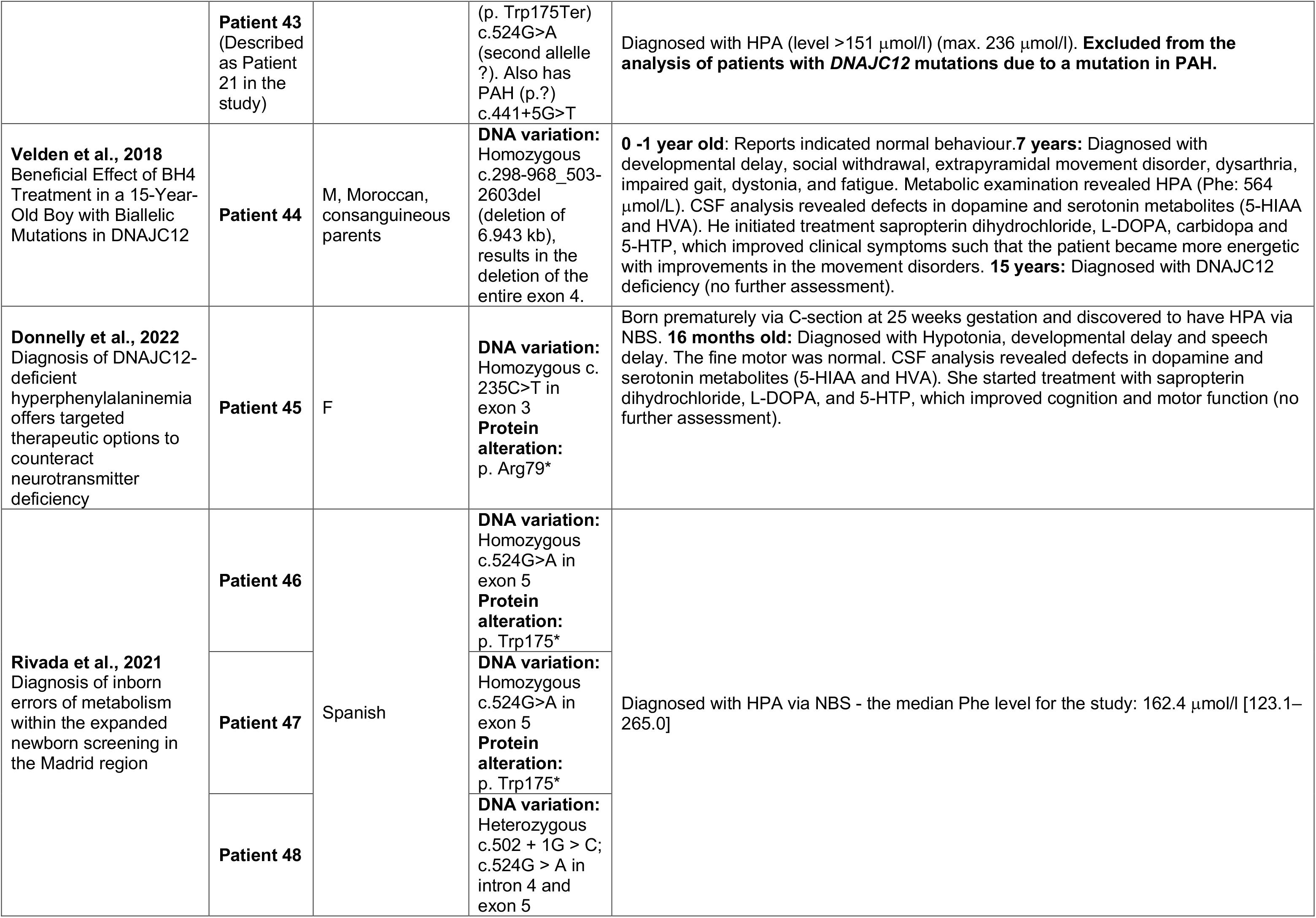

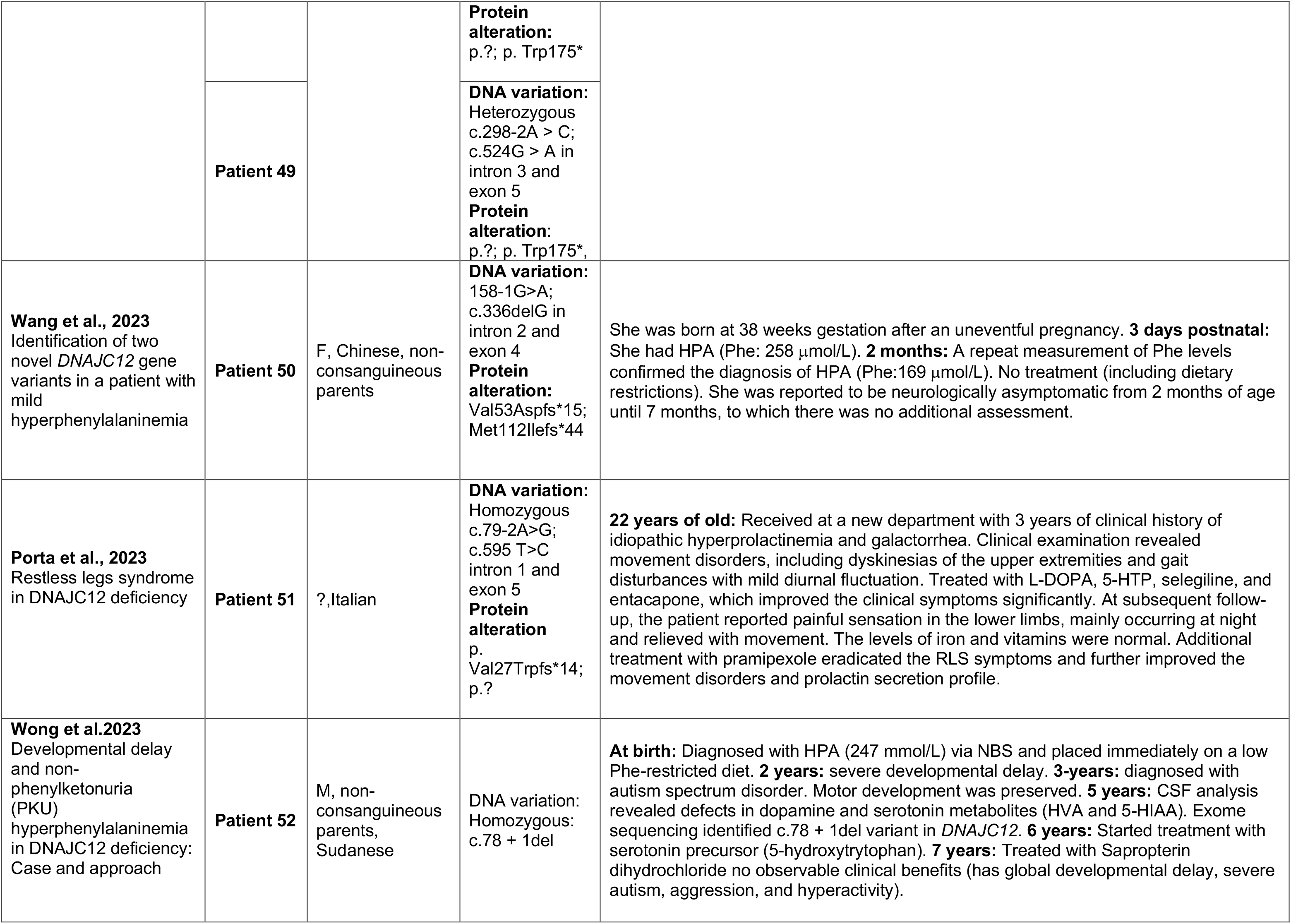

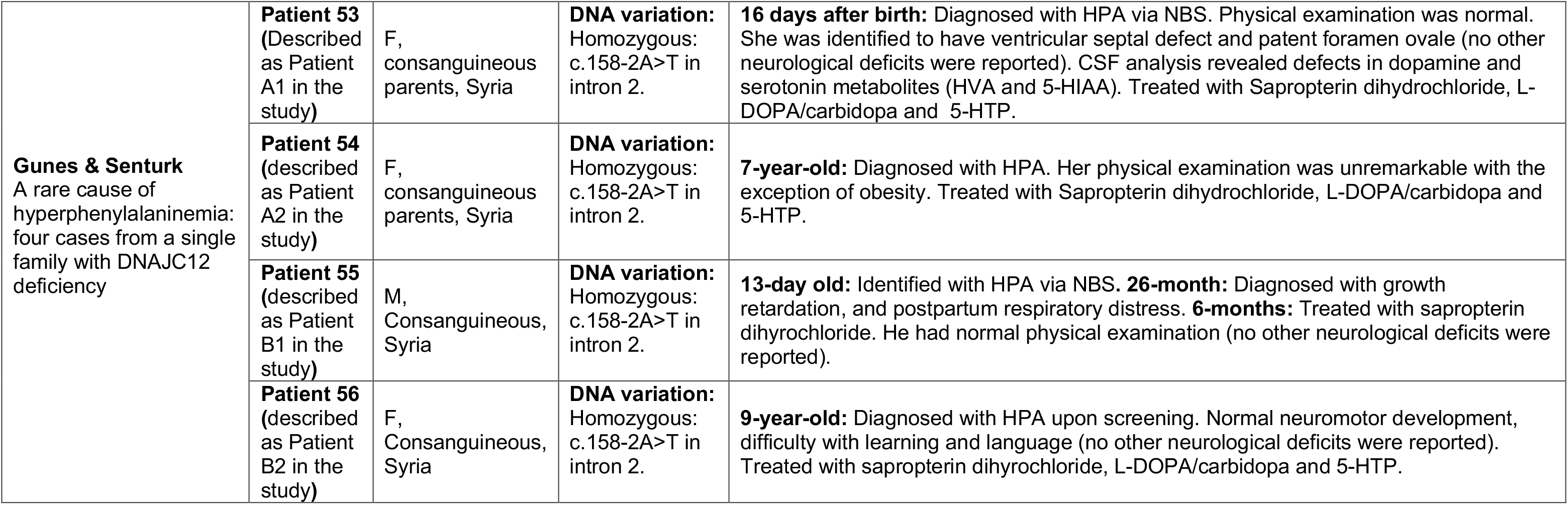

